# Middle aged carotid plaque and cognitive functions in later life: a population-based study

**DOI:** 10.64898/2026.03.23.26349039

**Authors:** Gisung Shin, Ali Tanweer Siddiquee, Min-Hee Lee, Yoonho Hwang, June Christoph Kang, Seungku Lee, Youngjin Kim, Bongjo Kim, Chol Shin, Nanhee Kim

## Abstract

**Background and Objectives:** Carotid plaque reflects systemic atherosclerosis and may serve as an early marker of cognitive decline, but its longitudinal association with cognitive trajectories remains unclear. We investigated whether carotid plaque status (small-to-medium size) was associated with cognitive performance over time in a population-based cohort.

**Methods:** This prospective cohort study followed-up neuropsychological assessment battery quadrennially in two cycles (2015-2018 and 2019-2022) from the baseline (2011-2014). Of 2,819 participants, 2,176 participants who were free of dementia and cerebrovascular disease at baseline with cognitive function testing at least two time points over the follow-up time were analyzed. Carotid plaque assessed by B-mode ultrasound sonography five segments were scanned in both left and right sides. The plaques were graded based on vessel thickness and the diameter of the lumen (none, small-to-medium, and large). We categorized our participants into without (none) and with the plaques (small-to-medium, and large combined) at baseline. The main outcomes were multivariable adjusted mean differences of cognitive test performances by baseline plaque status over time. The neuropsychological assessment battery included story recall, visual reproductions, verbal fluency, trail making, digit symbol – coding, and Stroop tests.

**Results:** Of the total, 291 (13.4%) participants had carotid plaque at baseline. There were no differences at baseline and 4-year. At 8-year follow-up, participants with carotid plaque performed significantly worse than participants without carotid plaque in visual reproduction delayed recall [mean difference −0.525 (95% CI: −0.915 to −0.135), p=.008], Stroop word reading [mean difference −2.732 (95% CI: −5.164 to −0.300), p=.028] and color reading [mean difference −3.573 (95% CI: −5.199 to −1.948), p<.001]. Additionally, participants with carotid plaque performed lower than those without carotid plaque on logical memory delayed recall [mean difference −1.577 (95% CI: −2.843 to −0.311), p=.015] at 8-year follow-up period.

**Discussion:** In this large cohort study, carotid plaque status at baseline was independently associated with in cognitive function decline, especially in non-verbal memory and executive functioning over 8-year follow-up period in the general population. Therefore, it may be important for earlier intervention on carotid plaque to preserve neurological health in middle-aged to older population.

## INTRODUCTION

Carotid plaque is a primary manifestation of atherosclerosis.^1^ It is a critical clinical marker to reflect both the local pathology in a particularly vulnerable arterial site and overall systemic burden of the disease.^2^ Intima-media thickness (IMT) also known to be a measurement for early-stage of atherosclerosis and had been extensively studied but its predictive value for cognitive decline remains highly contested due to longitudinal evidence.^3,4^ In contrast, carotid plaque is clinically more advanced predictor that substantially can contribute to long-term cardiovascular and cerebrovascular health risks.^3,5,6^

Previous studies^7–10^ have shown the association between IMT and cognitive functions yet the long-term relationship between carotid plaque and the cognitive decline over time remains to be fully elucidated. The beaver dam offspring study (BOSS) demonstrated the association between carotid plaque and cognitive function measured with both domain-specific tests (trail making test (TMT), grooved pegboard test (GPT)) and mini mental state examination (MMSE).^11^ While this cross sectional study initially found that higher carotid plaque scores were associated with lower performance on several cognitive tests, after adjusting for a comprehensive set of multivariable analysis, only a more severe plaque score (score 2) remained significantly associated with lower GPT and MMSE scores. Another cross-sectional study showed a moderate, but consistent, association between the presence of carotid plaques and poor cognitive performance in men for the MMSE and attention skill after adjusted for confounders.^12^ Although several large prospective population-based studies have investigated on cognitive outcomes in relation to carotid atherosclerosis, significant knowledge gap remains. Several studies were found to have been limited by the absence of baseline cognitive measurements and repeated measures with same participants over multiple time points, which may not fully capture the subtlety of continuous cognitive changes over time. ^3,10,13^ Therefore, our study aims to address the association between the carotid plaque status and cognitive function over an 8-year period.

## METHODS

### Study Participants

Korean Genome and Epidemiology Study (KoGES) is a population based study started in Ansan, South Korea Since the year 2001.^14^ We initiated additional core examinations including neuropsychological assessment battery for KoGES aging study (KoGES Ansan Aging Study) in 2011-2014, which was introduced in Exam 6 and Exam 7.^15^ Among 2,891 participants aged 49-79 who underwent ultrasonography at baseline, 2,726 participants completed neuropsychological examinations at both baseline (2011-2014) and follow-ups (2015-2018, and 2019-2022) after excluding those with baseline dementia (n=2) or cerebrovascular disease (n=91). We also further excluded participants who had incomplete neuropsychological tests (n=56), neuropsychological tests only performed once (n=448), and missing information in education, beck depression index (BDI), and low density lipoprotein cholesterol (LDL) (n=46). Finally, 2,176 participants who had neuropsychological assessments done at least 2 times (a total of 5,674 assessments) were included for analyses (Figure. 1).

**Figure 1.**
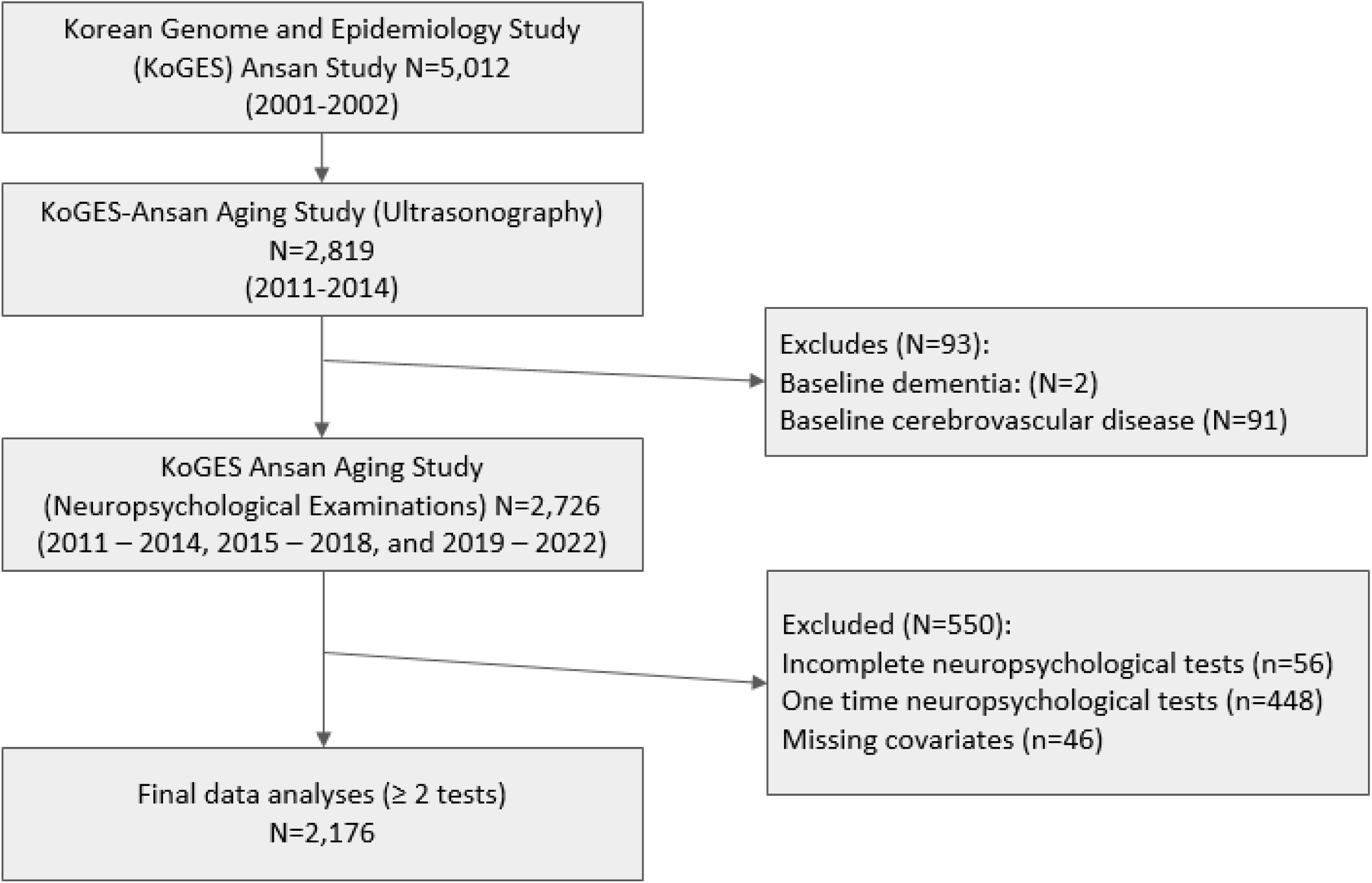
Flowchart of study participants from the KoGES Ansan Cohort Study. The flowchart shows the stepwise exclusion of participants based on no information on carotid plaque, history of neurological diseases, and absence/presence or complete/incomplete cognitive assessment data. 2,176 participants included in the final analysis.

This study was approved by the Institutional Review Board of Korea University Ansan Hospital (IRB NO 2006AS0045). All participants provided written informed consent and got agreements at the baseline and follow-up, in accordance with the Strengthening the Reporting of Observational Studies in Epidemiology (STROBE).

### Carotid Plaque Assessment

Carotid plaque presence was assessed using validated B-mode ultrasound sonography with a certified protocol (the University of Pittsburgh Ultrasound Research Lab) by a certified medical technician.^16^ Assessment from 2011 to 2012 used Sonosite Titan unit, while those from 2013 to 2014 used Sonosite MicroMaxx unit; both devices previously validated within KoGES.^17^ Plaque was defined as a distinct area of the vessel wall protruding into the lumen, with a thickness at least 50% greater than the adjacent IMT.^18^ Images were digitized and analyzed using M’ATH software (version 2.2, METRIS Co. Argenteuil, France). To assess the presence of plaque, five segments were scanned on each side left and right of the proximal and distal common carotid artery (CCA), the carotid bulb, internal carotid artery (ICA) and external carotid artery (ECA). Although carotid plaque was originally graded into four categories (grade 0: no observable plaque; grade 1: one small plaque, <30% of the vessel diameter; grade 2: one medium plaque, 30–50% of the vessel diameter or multiple small plaques; grade 3: one large plaque, >50% or multiple plaques with at least one medium plaque) according to the standard criteria^19^, grades 1 and 2 represent non-severe plaque categories and combined into small to medium plaque. For the analyses, plaque status was ultimately dichotomized into two groups: absence (none) and presence (small to medium or large) of plaques.

### Neuropsychological Assessment Battery

KoGES neuropsychological tests^20^ comprises the measures administered in the Framingham Heart Study (FHS) original and offspring cohorts, and its procedures and methods were adapted from the FHS protocol. Most tests were available in validated Korean versions but some were adapted from Wechsler Adult Intelligent Scale-Revised (WAIS-R) and Wechsler Memory Scale-III (WMS III). We evaluate verbal memory with story recall tests adapted from WMS-III. Verbal fluency was assessed using the phonemic subtests of the Controlled Oral Word Association Test and a category fluency task (animal naming). Visual memory was assessed with visual reproduction tests from the WAIS-R. Digit symbol – coding from the WAIS-IV was administered to assess visual processing and sustained attention. Simple attention was evaluated with the trail making test – A and Stroop test word reading. Executive function was measured with the stroop color reading task. This neuropsychological assessment battery emphasizes higher scores as better performance, except TMA.

### Covariates

This study participants of interviewees were administered questionnaires by well-trained interviewers. KOGES questionnaires included demographic, life-style, and history of disease status information (hypertension, diabetes, and cardiovascular disease) and biennially collected these data with a standardized manual. Alcohol consumption status asked the interviewee whether they had ever consumed the alcohol beverage or was there a time in their life when they regularly consumed or drank alcohol in past months. Self-reported smoking status was also collected as never, former, or current smokers. BDI score used to assess for measurements of the depressive symptom scores. Body mass index (BMI) was calculated as weight (kg) / height (m^2^). All blood samples were obtained in the morning after a 12-hour overnight fast and were immediately stored at −80°C for subsequent assays. Plasma concentrations of triglycerides, high-density lipoprotein cholesterol (HDL), and FBG levels enzymatically measured via 7600 Chemistry Analyzer (Hitachi; Tokyo, Japan). Apo-lipoprotein E (APOE) genotype was determined as part of a large-scale analysis using the Korea Biobank Array (KoreanChip), a microarray platform optimized for the Korean population containing over 833,000 genetic markers.^21^ APOE-ε4 carrier status defined as at least one ε4 carrier then considered as an ε4 carrier, else non-carrier. The white matter changes assessment is measured on a four-point age related white matter changes (ARWMC) scale (0=none, 1=focal, 2=beginning confluent, and 3=large confluent).^22^ Scores were assigned for five predefined regions (frontal, parieto-occipital, temporal, basal ganglia, and infratentorial) bilaterally. The total score was derived from summing the scores of these 10 regions, which results in a total score ranging from 0 to 30.

### Statistical Analysis

All statistical analyses were performed with SAS 9.4 software (SAS Institute, Cary, NC, USA). Baseline characteristics were analyzed with t-test for continuous variables, and χ^2^ test for categorical variables. Linear mixed model for repeated measurements with restricted maximum likelihood methods was used to examine the associations between carotid plaque status and cognitive performance over time. In the model, we account for unstructured covariance structure for within-person correlation across time. The model implicitly estimates missing values under a missing at random (MAR) assumption. We analyzed the mean differences (between participants without carotid plaque and with carotid plaque) and calculated the multivariable adjusted least squares means with 95% confidence intervals of the cognitive performance of carotid plaque groups to compare significant differences between the groups over time (at baseline, 4-year, and 8-year follow-up prospectively). Covariates were adjusted for baseline age, sex, education, drinking, smoking, HDL, LDL, hypertension (HTN), diabetes (DM), cardiovascular disease (CVD), and BDI score. To test the robustness of our findings, we performed sensitivity analyses to repeat the primary analysis further adjusting for ARWMC and for APOE-ε4 carrier status in separate models. ARWMC reduced the blood flow from small vessel disease to disrupt wiring connectivity, which can lead executive dysfunction.^23^ While APOE-ε4 carrier status impaired clearance of amyloid beta proteins to buildup waste, which may impair memory functions.^24^ A two tailed p-value <0.05 was considered as statistically significant.

## RESULTS

### Baseline Characteristics

The baseline characteristics of study participants by carotid plaque status are shown in Table 1. Participants with carotid plaque were about 3 years older than without carotid plaque group (p <.001). There were more men in participants with carotid plaque and higher education level was proportionally lower in with carotid plaque group compared to without carotid plaque (p<0.01). For the participants with carotid plaque, past smokers and current smokers were 37.8% and 17.9%, respectively. In contrast, those who without carotid plaque was predominantly never smokers (64.8%). For the presence of carotid plaque groups, past drinkers and current drinkers were 9.3% and 52.6%, respectively. On the other hand, those who absence of carotid plaque group as never drinkers were 50.6%. In overall, 912 participants had HTN (41.9%), 638 participants had DM (29.3%), and 112 participants had CVD (5.1%). Each of these conditions was significantly more prevalent among participants with carotid plaque compared to those without carotid plaque: HTN (59.5% vs 39.2%), DM (40.2% vs 27.6%), except CVD (6.9% vs 4.9%) (Table 1). On the other hand, BMI, BDI scores, HDL and LDL cholesterols and regular exercise status were not significant between the carotid plaque groups.

**Table 1.**
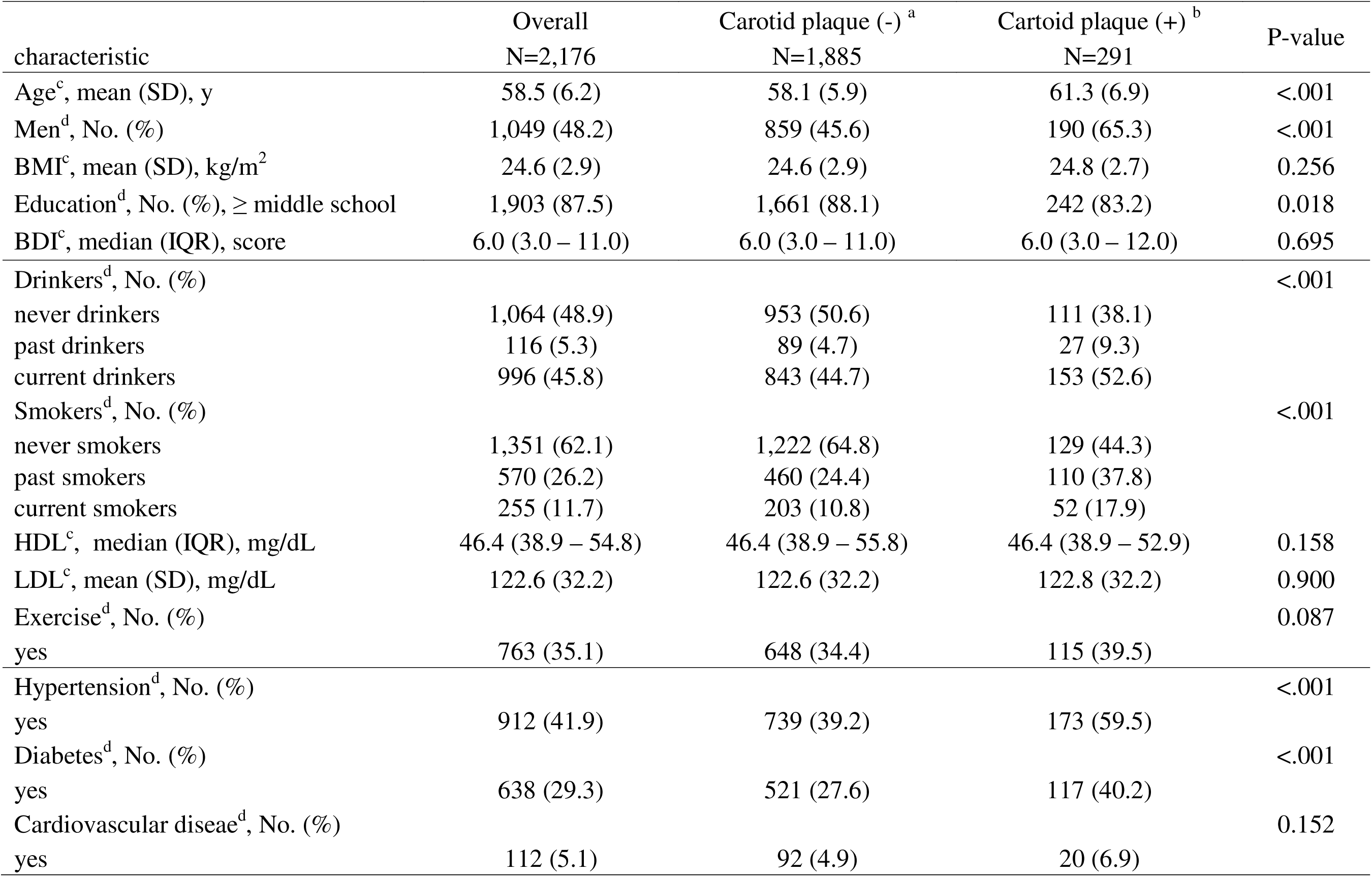
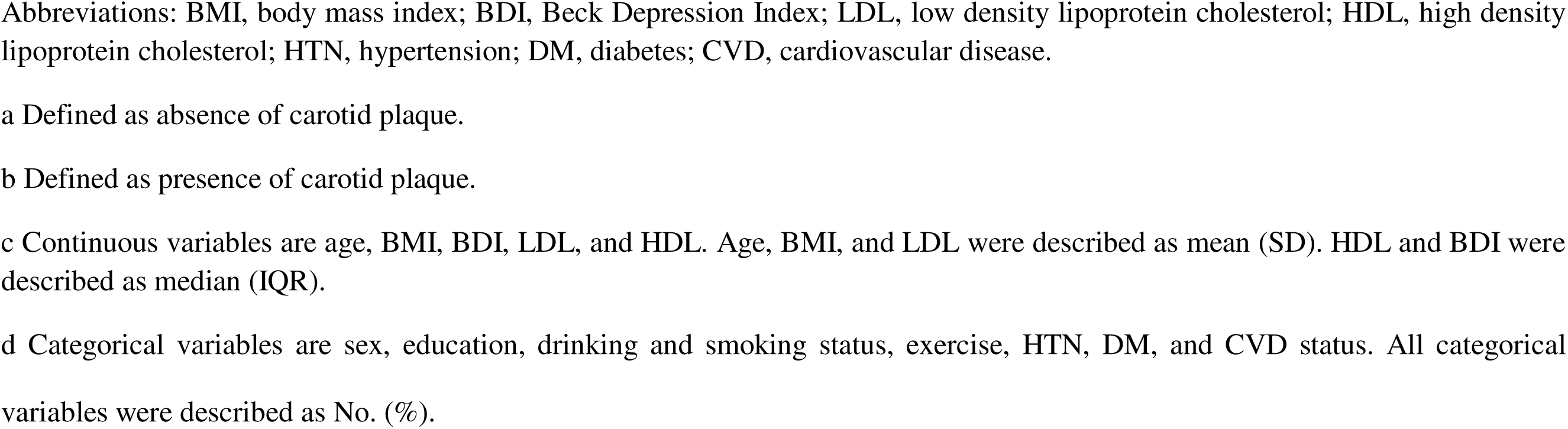
Baseline general characteristics of the study participants (N=2,176) by carotid plaque status.

| characteristic | Overall<br>N=2,176 | Carotid plaque (-) <sup>a</sup><br>N=1,885 | Cartoid plaque (+) <sup>b</sup><br>N=291 | P-value |
| --- | --- | --- | --- | --- |
| Age<sup>c</sup>, mean (SD), y | 58.5 (6.2) | 58.1 (5.9) | 61.3 (6.9) | <.001 |
| Men<sup>d</sup>, No. (%) | 1,049 (48.2) | 859 (45.6) | 190 (65.3) | <.001 |
| BMI<sup>c</sup>, mean (SD), kg/m<sup>2</sup> | 24.6 (2.9) | 24.6 (2.9) | 24.8 (2.7) | 0.256 |
| Education<sup>d</sup>, No. (%), ≥ middle school | 1,903 (87.5) | 1,661 (88.1) | 242 (83.2) | 0.018 |
| BDI<sup>c</sup>, median (IQR), score | 6.0 (3.0 – 11.0) | 6.0 (3.0 – 11.0) | 6.0 (3.0 – 12.0) | 0.695 |
| Drinkers<sup>d</sup>, No. (%) |  |  |  | <.001 |
| never drinkers | 1,064 (48.9) | 953 (50.6) | 111 (38.1) |  |
| past drinkers | 116 (5.3) | 89 (4.7) | 27 (9.3) |  |
| current drinkers | 996 (45.8) | 843 (44.7) | 153 (52.6) |  |
| Smokers<sup>d</sup>, No. (%) |  |  |  | <.001 |
| never smokers | 1,351 (62.1) | 1,222 (64.8) | 129 (44.3) |  |
| past smokers | 570 (26.2) | 460 (24.4) | 110 (37.8) |  |
| current smokers | 255 (11.7) | 203 (10.8) | 52 (17.9) |  |
| HDL<sup>c</sup>, median (IQR), mg/dL | 46.4 (38.9 – 54.8) | 46.4 (38.9 – 55.8) | 46.4 (38.9 – 52.9) | 0.158 |
| LDL<sup>c</sup>, mean (SD), mg/dL | 122.6 (32.2) | 122.6 (32.2) | 122.8 (32.2) | 0.900 |
| Exercise<sup>d</sup>, No. (%) |  |  |  | 0.087 |
| yes | 763 (35.1) | 648 (34.4) | 115 (39.5) |  |
| Hypertension<sup>d</sup>, No. (%) |  |  |  | <.001 |
| yes | 912 (41.9) | 739 (39.2) | 173 (59.5) |  |
| Diabetes<sup>d</sup>, No. (%) |  |  |  | <.001 |
| yes | 638 (29.3) | 521 (27.6) | 117 (40.2) |  |
| Cardiovascular disease<sup>d</sup>, No. (%) |  |  |  | 0.152 |
| yes | 112 (5.1) | 92 (4.9) | 20 (6.9) |  |

Abbreviations: BMI, body mass index; BDI, Beck Depression Index; LDL, low density lipoprotein cholesterol; HDL, high density lipoprotein cholesterol; HTN, hypertension; DM, diabetes; CVD, cardiovascular disease.
a Defined as absence of carotid plaque.
b Defined as presence of carotid plaque.
c Continuous variables are age, BMI, BDI, LDL, and HDL. Age, BMI, and LDL were described as mean (SD). HDL and BDI were described as median (IQR).
d Categorical variables are sex, education, drinking and smoking status, exercise, HTN, DM, and CVD status. All categorical variables were described as No. (%).

### Distribution of carotid plaque grade

The prevalence and distribution of carotid plaque by plaque grade is shown in Table 2. Of 291 participants with carotid plaque, the majority of the participants were small to medium sized plaque (n=283; 13.0%) and the rest were large plaque (n=8; 0.37%). Indeed, a total of 2,176 participants, the majority participants of this study did not have carotid plaque (86.6%) at baseline.

**Table 2.** The prevalence and distribution of carotid plaque status by plaque grades (N=2,176).

| **plaque status** | **Plaque Grade** | **Number of participants (n)** | **percentages (%)** |
| --- | --- | --- | --- |
| Absence of carotid plaque | no observable plaque | 1,885 | 86.6 |
| Presence of carotid plaque | small to medium plaque ($\leq 50\%$ of the vessel<sup>a</sup> diameter) | 283 | 13.0 |
| | large plaque ($> 50\%$ of the vessel<sup>a</sup> diameter) | 8 | 0.37 |
| Total | Overall Plaque | 2,176 | 100 |

<sup>a</sup> Vessel denotes the local lumen diameter at the scanned carotid segment (proximal and distal CCA, carotid bulb, ICA, and ECA). Grades indicate plaque size relative to lumen diameter.

### Changes in cognitive functions in relation to carotid plaque status

Of the 12 neuropsychological tests, none of the scores significantly differ at baseline and 4-year follow-up (Figure 2, Table 3). However, logical memory [mean difference −1.577 (95% CI: - 2.843 to −0.311), p=.015] and visual reproduction delayed recall [mean difference −0.525 (95% CI: −0.915 to −0.135), p=.008], and stroop word reading [mean difference −2.732 (95% CI: −5.164 to −0.300), p=0.028] and color reading tests [mean difference −3.573 (95% CI: −5.199 to −1.948), p<.001] showed lower performance in participants with carotid plaque compared to without carotid plaque group over an 8-year period (Figure 2, Table 3). The logical memory and visual reproduction immediate recall and recognition, verbal fluency letter and animal, trail making test A, and digit symbol-coding tests scores were not significantly different between carotid plaque groups (Figure 2).

**Figure 2.**
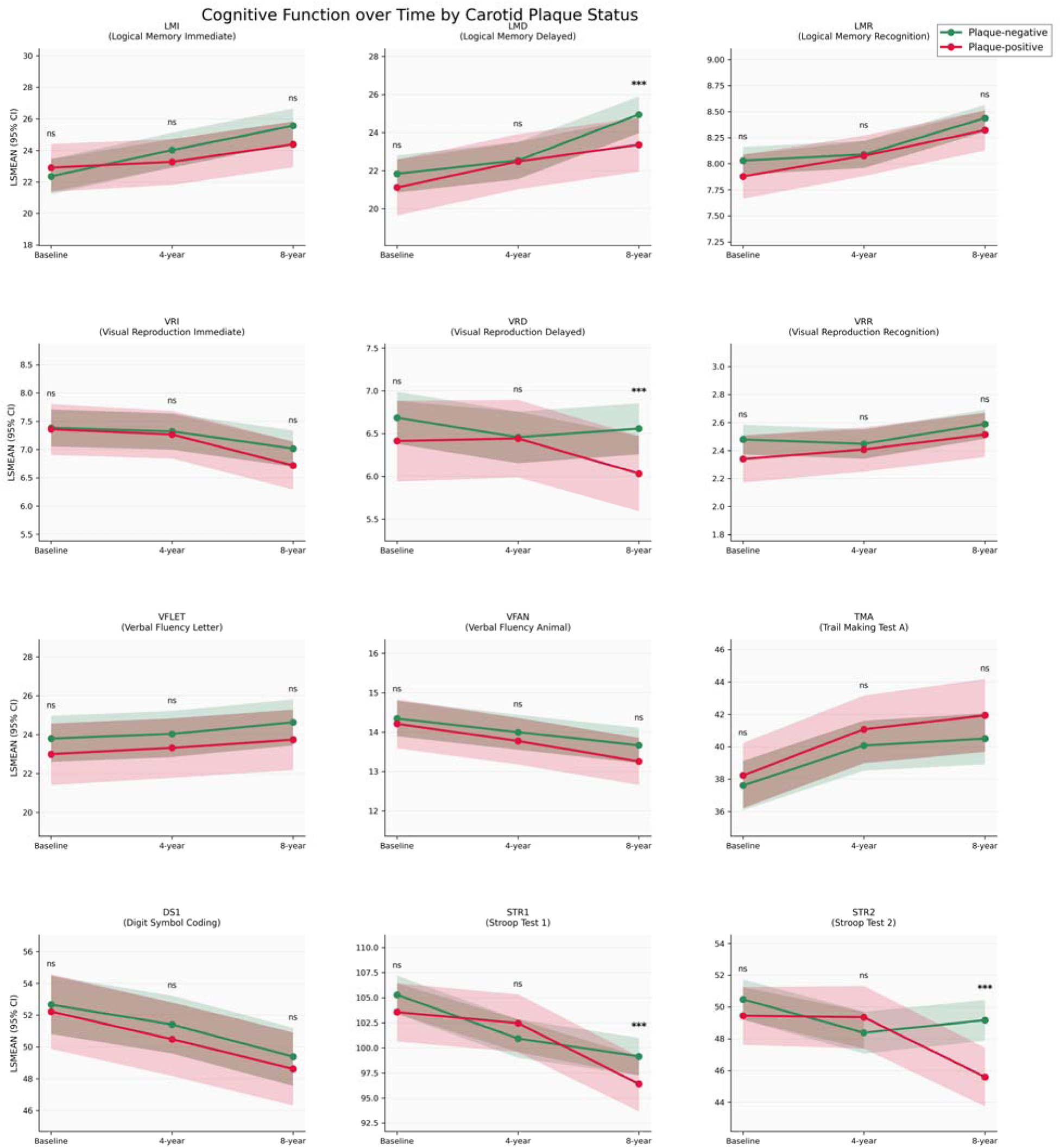
Associations of carotid plaque status and cognitive functions over time. Linear mixed models multivariable adjusted mean differences for repeated measurements are plotted. Story recall and verbal fluency letter test increased over time both groups in carotid plaque. Stroop tests declined over time in the participants with carotid plaque compared to the participants without carotid plaque. Notes: ns = non-significant, *** p < 0.05 = significant. lines with surrounded colored represent 95% confidence intervals.

**Table 3.** Multivariable adjusted^a^ mean difference (95% Cl) of cognitive function test scores using linear mixed models for repeated measurements between the carotid plaque status groups overtime (N = 2,176).

|  |  | Baseline |  | 4-year |  | 8-year |  |
| --- | --- | --- | --- | --- | --- | --- | --- |
|  |  | Estimate (95% CI) | p-value | Estimate (95% CI) | p-value | Estimate (95% CI) | p-value |
| **Story recall tests** |  |  |  |  |  |  |  |
|  | Immediate recall | 0.555 (-0.708, 1.817) | 0.389 | -0.751 (-1.930, 0.429) | 0.212 | -1.176 (-2.358, 0.006) | 0.051 |
|  | Delayed recall | -0.714 (-2.041, 0.613) | 0.292 | -0.062 (-1.352, 1.229) | 0.926 | **-1.577 (-2.843, -0.311)** | **0.015** |
|  | Recognition | -0.152 (-0.348, 0.043) | 0.127 | -0.013 (-0.188, 0.162) | 0.883 | -0.116 (-0.289, 0.057) | 0.188 |
| **Visual Reproduction tests** |  |  |  |  |  |  |  |
|  | Immediate recall | -0.025 (-0.406, 0.355) | 0.897 | -0.056 (-0.398, 0.285) | 0.747 | -0.299 (-0.646, 0.047) | 0.091 |
|  | Delayed recall | -0.273 (-0.705, 0.159) | 0.215 | -0.014 (-0.422, 0.394) | 0.946 | **-0.525 (-0.915, -0.135)** | **0.008** |
|  | Recognition | -0.139 (-0.294, 0.016) | 0.079 | -0.040 (-0.183, 0.103) | 0.584 | -0.075 (-0.219, 0.069) | 0.308 |
| **Verbal Fluency Tests** |  |  |  |  |  |  |  |
|  | Phonemic | -0.799 (-2.094, 0.496) | 0.226 | -0.725 (-1.958, 0.509) | 0.249 | -0.898 (-2.154, 0.358) | 0.161 |
|  | Category | -0.137 (-0.663, 0.390) | 0.611 | -0.225 (-0.711, 0.262) | 0.366 | -0.409 (-0.903, 0.086) | 0.105 |
| **Trail making Test** |  |  |  |  |  |  |  |
|  | Trails A time | 0.618 (-1.018, 2.255) | 0.459 | 0.999 (-0.752, 2.749) | 0.263 | 1.446 (-0.534, 3.425) | 0.152 |
| **Digit symbol test** |  |  |  |  |  |  |  |
|  | Coding | -0.434 (-2.294, 1.426) | 0.647 | -0.918 (-2.715, 0.879) | 0.316 | -0.769 (-2.559, 1.021) | 0.400 |
| **Stroop tests** |  |  |  |  |  |  |  |
|  | Word reading | -1.733 (-4.360, 0.895) | 0.196 | 1.533 (-1.076, 4.141) | 0.249 | **-2.732 (-5.164, -0.300)** | **0.028** |
|  | Color reading | -1.025 (-2.604, 0.555) | 0.204 | 0.970 (-0.822, 2.762) | 0.289 | **-3.573 (-5.199, -1.948)** | **<.001** |

Abbreviations: LMI, logical memory immediate recall; LMD, logical memory delayed recall; LMR, logical memory recognition; VRI, visual reproduction immediate recall; VRD, visual reproduction delayed recall; VRR, visual reproduction recognition; VFLET, verbal fluency – letter test; VFAN, verbal fluency – animal test; TMA, trail making test A; and DS – coding, digit symbol – coding.
a Adjusted Model: Multi-variable adjusted age, sex, education, bmi, smoking, drinking, HDL, LDL, hypertension, diabetes, and cardiovascular disease, BDI score.

### Sensitivity Analyses

The first sensitivity analysis was done by further adjust for baseline ARWMC (n=2,163) to reflect small vessel disease burden (Supplement Table 1). Compared to participants without carotid plaque, logical memory and visual reproduction delayed recall [mean difference −1.578 (95% CI:-2.845 to −0.311), p=.015] and [mean difference −0.533 (95% CI:-0.924 to −0.143), p=.007], and Stroop word reading [mean difference −2.816 (95% CI:-5.251 to −0.381), p=0.023] and color reading [mean difference −3.646 (95% CI:-5.276 to −2.016), p<.001] tests scores were still remain significantly performed lower in those with carotid plaque after further adjusted ARWMC at 8-year follow-up (Supplement Table 1).

Second sensitivity analysis adjusted for baseline APOE-ε4 carrier status (n=1,842) to reflect major genetic risk factor for dementia (Supplement Table 2). Visual reproduction [mean difference −0.475 (95% CI: −0.905 to −0.046), p=0.030], and stroop color reading [mean difference −3.366 (95% CI: −5.041 to −1.692), p<.001] tests were remaining significant but logical memory delayed recall and stroop word reading tests attenuated after further adjusted for APOE-ε4 carrier status (Supplement Table 2). Our results remained robust and supported our primary findings, which is not driven by underlying small vessel pathology or genetic susceptibility.

## DISCUSSION

In this large population-based prospective cohort study, the presence of carotid plaques, especially small to medium sized, was independently associated with cognitive decline in logical memory, visual reproduction delayed recall and stroop tests over 8-year follow-up period. To the best of our knowledge, this is the first time to investigate the relationship between carotid plaque status and cognitive functions over time in middle-aged to older population based cohort study with repeated measurements.

In previous few population-based prospective studies, similar atherosclerosis-related cognitive declines were reported. Arntzen, et al (2012) examined associations between plaque status and cognitive performance in 3 standardized cognitive tests of word memory, digit-symbol (WAIS), and taping tests and reported the presence of carotid plaques significantly associated with lower performance on verbal memory test after multivariable adjusted for age, sex, education, depression, and vascular risk factors in stroke free participants for 7-years follow up.^3^ Lin, et al (2020) also reported both functional (carotid stiffness) and structural (cIMT and carotid plaque) atherosclerosis markers associated with MoCA (executive and language index) score after 10 years with stroke, myocardial infarction, and dementia free participants adjusted for age, sex, education, HTN, DM, hypercholesterolemia, and smoking status.^13^ Unlike these previous studies, our multivariable-adjusted analyses showed no significant differences in cognitive performance between participants with or without carotid plaque at baseline and 4-year follow-up, after excluding participants with neurological disease (dementia and cerebrovascular disease) at baseline (Figure 2, Table 3). However, participants with carotid plaques were significantly associated with lower cognitive performance in logical memory and visual reproduction delayed recall and stroop tests at 8-year follow up (Figure 2, Table 3).

On the other hand, few previous studies reported negative results for cognitive functions in relation to carotid plaque status. Northern Manhattan Study (mean age of 70 years old) reported carotid plaque status or area was not associated with cognitive decline.^9^ The epidemiology of hearing loss study (EHLS) in Beaver Dam (mean age of 66.8 years old) also did not find associations between carotid plaque status and cognitive tests 10 years later.^10^ Unlike these studies, we had a relatively healthy younger (mean age of 58.5 years old) population to investigate on associations between carotid plaque and cognitive performance over 8-year period with large middle-aged to older population. Carotid plaque may impair cognition through hypo-perfusion^25^ and embolic pathways^26^, which can lead to structural changes such as white matter hyper-intensities^27^ and cortical atrophy^28^. These biological rationales for a direct link between the presence of carotid plaque and cognitive decline. In our study, participants with carotid plaque demonstrated a deficit in executive functions (stroop color reading tests) compared to those without carotid plaque. Additionally, simple attention measures showed same-direction trends that did not uniformly reach significance. Notably, visual reproduction delayed recall performance was significantly lower in participants with carotid plaque than those without carotid plaque even after further adjustment for ARWMC. This suggested the detrimental effects of carotid plaque on those cognitive domains are not solely mediated by the structural brain changes or vascular pathway (Supplement Table 1). We also separately adjusted APOE-ε4 carrier status to distinguish this vascular pathway from genetic predisposition and still remained significant except logical memory delayed recall and stroop word reading tests possibly due to smaller sample size in the analysis (Supplement Table 2). However, logical memory functions increased over time regardless of carotid plaque status may reflect learning effects with more familiar with test items, concepts and strategies.^29,30^ The WMS logical memory is particularly sensitive to learning effects, as individuals often recall the stimulus stories at subsequent repeated assessments.^29,31^ In our study, we used a short story adapted to the Korean context that contains negative emotional content. Although practice-related gains may have typically greatest at short retest intervals due to prior studies have reported those learning effects may persist about 3 to 7 year or even longer.^32,33^ A plausible explanation for this durability is the context-rich and emotionally salient narratives may be encoded similarly to a personal experience events.

This study has several limitations. First, this prospective cohort study was unable to evaluate a dose-response relationship between carotid plaque and cognitive performance due to lack of differentiation of small to medium sized carotid plaques. Second, our study was not designed to test learning effects, which cannot completely be ruled out. Nonetheless, participants with carotid plaque generally had lower cognitive scores compared to those without carotid plaque, suggesting that carotid plaque group may fail to learn (Figure 2). Third, the analysis of multiple cognitive assessments inflates the risk of type 1 error (a false positive) due to multiple comparison. However, we refrained from applying formal statistical correction because they often increase the risk of type 2 error (a false negative) while protecting against type 1 error.^34^ This could potentially cause a true effect to be overlooked. Fourth, we had excluded participants from analysis due to incomplete data such as missing covariates or lack of repeated measurements on neuropsychological assessments (Figure 1). Although our populations are reasonably large to compare general characteristics of included and excluded participants. Compared to included participants, excluded participants were significantly older, less educated, had more comorbidities (HTN and DM) (Supplement Table 3). The retention of a relatively younger and healthier study population creates a potential selection bias on underestimation of our finding for cognitive decline due to a ‘healthy cohort effect’. Participants, enrolled since 2001, have had regular access to the Korean national health system, which provides biennial health check-ups. This likely increased their awareness and management of conditions like HTN, potentially limiting the applicability of our results to populations in healthcare settings with fewer resources.

In conclusion, our study findings support the presence of small to medium sized carotid plaque may be an independent risk factor for cognitive decline, particularly in executive functions. This finding adds significant evidence to the limited literature linking asymptomatic atherosclerosis to future cognitive decline. Therefore, the early prevention and managements of carotid plaque could be essential strategy to preserve neurological health in normally aging population. Furthermore, future research should focus on more precise quantification of carotid plaque characteristics combined with longer follow-up periods in the middle to older aged populations, to improve the prediction of long-term cognitive outcomes.

## Data Availability

All data produced in the present work are contained in the manuscript

## Author contributions

our corresponding authors had full access to all of the data in the study and took responsibility for the integrity of data and the accuracy of the data analysis.

Concept and design: G. Shin, A Siddiquee, M. Lee, June Kang, N. Kim, C. Shin.

Acquisition, analysis, or interpretation data: G. Shin, A Siddiquee, Y. Hwang, Y. Kim, B. Kim, S. Lee, C. Shin, N. Kim.

Drafting the manuscript: G. Shin.

Critical revision of manuscript for important intellectual content: All authors.

Statistical Analysis: G. Shin.

Obtained funding: N. Kim.

Administrative, technical, or material support: G. Shin, Y. Hwang, S. Lee, C. Shin, and N. Kim.

## Role of the funder or sponsor

The funders had no role in the design and conduct of the study; collection, management, analysis, and interpretation of the data; preparation, review, or approval of the manuscript; and decision to submit the manuscript for publication.

## Acknowledgements

This study was performed with bio-resources from National Biobank of Korea, the National Institute of Health (NIH), Republic of Korea. Genotype data were provided by the Collaborative Genome Program for Fostering New Post-Genome Industry (3000-3031b). Gahyun Oh also contributed her time and effort to make power point of this study for internal presentation before getting approval. AIFFEL at MODULAB also support this study.

