## Supplement Tables for "Middle aged carotid plaque and cognitive functions in later life: a population-based study"

Supplement Table 1. Multivariable and ARWMC adjusted mean difference (95% Cl) of cognitive function test scores using linear mixed models for repeated measurements between the carotid plaque status groups overtime (N = 2,163).

Supplement Table 2. Multivariable and ApoE4 adjusted mean difference (95% Cl) of cognitive function test scores using linear mixed models for repeated measurements between the carotid plaque status groups overtime (N = 1,842).

Supplement Table 3. Baseline Characteristics comparison between included (N= 2,176) and excluded (N=550) study participants.

Supplement Table 1. Multivariable and ARWMC adjusted^a^ mean difference (95% Cl) of cognitive function test scores using linear mixed models for repeated measurements between the carotid plaque status groups overtime (N = 2,163).

|  |  | **Baseline** | | **4-year** | | **8-year** | |
| --- | --- | --- | --- | --- | --- | --- | --- |
|  |  | Estimate (95% CI) | p-value | Estimate (95% CI) | p-value | Estimate (95% CI) | p-value |
| **Story recall tests** |  |  |  |  |  |  |  |
|  | Immediate recall | 0.589 (-0.675, 1.852) | 0.361 | -0.718 (-1.900, 0.464) | 0.234 | -1.154 (-2.338, 0.031) | 0.056 |
|  | Delayed recall | -0.704 (-2.030, 0.623) | 0.298 | -0.060 (-1.351, 1.231) | 0.928 | **-1.578 (-2.845, -0.311)** | **0.015** |
|  | Recognition | -0.149 (-0.345, 0.047) | 0.136 | -0.010 (-0.185, 0.165) | 0.913 | -0.114 (-0.287, 0.059) | 0.197 |
| **Visual Reproduction tests** |  |  |  |  |  |  |  |
|  | Immediate recall | -0.022 (-0.403, 0.360) | 0.912 | -0.056 (-0.398, 0.287) | 0.749 | -0.297 (-0.645, 0.051) | 0.094 |
|  | Delayed recall | -0.286 (-0.718, 0.147) | 0.195 | -0.011 (-0.419, 0.398) | 0.958 | **-0.533 (-0.924, -0.143)** | **0.007** |
|  | Recognition | -0.138 (-0.294, 0.017) | 0.080 | -0.037 (-0.180, 0.106) | 0.610 | -0.078 (-0.222, 0.066) | 0.290 |
| **Verbal Fluency Tests** |  |  |  |  |  |  |  |
|  | Phonemic | -0.776 (-2.074, 0.523) | 0.242 | -0.659 (-1.895, 0.577) | 0.296 | -0.862 (-2.120, 0.397) | 0.179 |
|  | Category | -0.109 (-0.636, 0.418) | 0.685 | -0.198 (-0.685, 0.290) | 0.426 | -0.394 (-0.890, 0.102) | 0.119 |
| **Trail making Test** |  |  |  |  |  |  |  |
|  | Trails A time | 0.595 (-1.046, 2.235) | 0.477 | 0.942 (-0.813, 2.697) | 0.293 | 1.398 (-0.586, 3.382) | 0.167 |
| **Digit symbol test** |  |  |  |  |  |  |  |
|  | Coding | -0.502 (-2.363, 1.359) | 0.597 | -0.961 (-2.760, 0.838) | 0.295 | -0.824 (-2.616, 0.967) | 0.367 |
| **Stroop tests** |  |  |  |  |  |  |  |
|  | Word reading | -1.828 (-4.458, 0.802) | 0.173 | 1.459 (-1.149, 4.067) | 0.273 | **-2.816 (-5.251, -0.381)** | **0.023** |
|  | Color reading | -1.085 (-2.668, 0.499) | 0.179 | 0.930 (-0.867, 2.726) | 0.310 | **-3.646 (-5.276, -2.016)** | **<.001** |

Abbreviations: LMI, logical memory immediate recall; LMD, logical memory delayed recall; LMR, logical memory recognition; VRI, visual reproduction immediate recall; VRD, visual reproduction delayed recall; VRR, visual reproduction recognition; VFLET, verbal fluency – letter test; VFAN, verbal fluency – animal test; TMA, trail making test A; and DS – coding, digit symbol – coding.

a Adjusted Model: Multi-variable adjusted age, sex, education, bmi, smoking, drinking, HDL, LDL, hypertension, diabetes, and cardiovascular disease, BDI score and ARWMC.

Supplement Table 2. Multivariable and ApoE4 adjusted^a^ mean difference (95% Cl) of cognitive function test scores using linear mixed models for repeated measurements between the carotid plaque status groups overtime (N = 1,842).

|  |  | **Baseline** | | **4-year** | | **8-year** | |
| --- | --- | --- | --- | --- | --- | --- | --- |
|  |  | Estimate (95% CI) | p-value | Estimate (95% CI) | p-value | Estimate (95% CI) | p-value |
| **Story recall tests** |  |  |  |  |  |  |  |
|  | Immediate recall | 0.574 (-0.805, 1.954) | 0.414 | -0.335 (-1.617, 0.946) | 0.608 | -1.011 (-2.296, 0.274) | 0.123 |
|  | Delayed recall | -0.606 (-2.057, 0.845) | 0.412 | -0.181 (-1.587, 1.225) | 0.801 | -1.214 (-2.595, 0.168) | 0.085 |
|  | Recognition | -0.133 (-0.345, 0.079) | 0.221 | 0.013 (-0.177, 0.203) | 0.893 | -0.076 (-0.265, 0.112) | 0.427 |
| **Visual Reproduction tests** |  |  |  |  |  |  |  |
|  | Immediate recall | 0.075 (-0.342, 0.492) | 0.724 | 0.019 (-0.354, 0.392) | 0.920 | -0.310 (-0.690, 0.069) | 0.109 |
|  | Delayed recall | -0.212 (-0.686, 0.262) | 0.379 | -0.099 (-0.549, 0.349) | 0.663 | **-0.475 (-0.905, -0.046)** | **0.030** |
|  | Recognition | -0.106 (-0.273, 0.062) | 0.216 | -0.078 (-0.235, 0.078) | 0.325 | -0.051 (-0.209, 0.106) | 0.523 |
| **Verbal Fluency Tests** |  |  |  |  |  |  |  |
|  | Phonemic | -0.203 (-1.611, 1.204) | 0.777 | -0.243 (-1.592, 1.106) | 0.724 | -0.310 (-1.692, 1.072) | 0.660 |
|  | Category | -0.025 (-0.596, 0.547) | 0.933 | -0.148 (-0.678, 0.382) | 0.583 | -0.239 (-0.787, 0.307) | 0.390 |
| **Trail making Test** |  |  |  |  |  |  |  |
|  | Trails A time | 0.549 (-1.202, 2.301) | 0.539 | 0.251 (-1.645, 2.147) | 0.795 | 0.808 (-1.399, 3.016) | 0.473 |
| **Digit symbol test** |  |  |  |  |  |  |  |
|  | Coding | 0.427 (-1.607, 2.461) | 0.681 | 0.062 (-1.897, 2.020) | 0.951 | 0.257 (-1.690, 2.205) | 0.796 |
| **Stroop tests** |  |  |  |  |  |  |  |
|  | Word reading | -1.497 (-4.349, 1.355) | 0.304 | 1.898 (-0.925, 4.720) | 0.188 | -2.131 (-4.758, 0.497) | 0.112 |
|  | Color reading | -1.021 (-2.692, 0.651) | 0.223 | 0.601 (-1.121, 2.322) | 0.494 | **-3.366 (-5.041, -1.692)** | **<.001** |

Abbreviations: LMI, logical memory immediate recall; LMD, logical memory delayed recall; LMR, logical memory recognition; VRI, visual reproduction immediate recall; VRD, visual reproduction delayed recall; VRR, visual reproduction recognition; VFLET, verbal fluency – letter test; VFAN, verbal fluency – animal test; TMA, trail making test A; and DS – coding, digit symbol – coding.

a Adjusted Model: Multi-variable adjusted age, sex, education, bmi, smoking, drinking, HDL, LDL, hypertension, diabetes, and cardiovascular disease, BDI score and APOE4.

Supplement Table 3. Baseline Characteristics comparison between included (N= 2,176) and excluded (N=550) study participants.

|  | Included^a^ | Excluded^b^ | P-value |
| --- | --- | --- | --- |
| characteristic | N=2,176 | N=550 |  |
| Age^c^, mean (SD), y | 58.5 (6.2) | 61.5 (8.5) | <.001 |
| Men^d^, No. (%) | 1,049 (48.2) | 259 (47.1) | 0.640 |
| BMI^c^, mean (SD), kg/m^2^ | 24.6 (2.9) | 24.6 (3.0) | 0.912 |
| Education^d^, No. (%), ≥ middle school (N=2,724) | 1,903 (87.5) | 417 (76.1) | <.001 |
| BDI^c^, median (IQR), score | 6.0 (3.0 – 11.0) | 6.0 (3.0 - 12.0) | 0.305 |
| Drinkers^d^, No. (%) |  |  | 0.177 |
| never drinkers | 1,064 (48.9) | 288 (52.4) |  |
| past drinkers | 116 (5.3) | 34 (6.2) |  |
| current drinkers | 996 (45.8) | 228 (41.5) |  |
| Smokers^d^, No. (%) |  |  | 0.741 |
| never smokers | 1,351 (62.1) | 338 (61.5) |  |
| past smokers | 570 (26.2) | 141 (25.6) |  |
| current smokers | 255 (11.7) | 71 (12.9) |  |
| HDL^c^, median (IQR), mg/dL | 46.4 (38.9 – 54.8) | 46.4 (38.0 - 54.8) | 0.118 |
| LDL^c^, mean (SD), mg/dL | 122.6 (32.2) | 121.5 (32.6) | 0.472 |
| Exercise^d^, No. (%) |  |  | 0.295 |
| yes | 763 (35.1) | 206 (37.5) |  |
| Hypertension^d^, No. (%) |  |  | <.001 |
| yes | 912 (41.9) | 293 (53.3) |  |
| Diabetes^d^, No. (%) |  |  | 0.003 |
| yes | 638 (29.3) | 197 (35.8) |  |
| Cardiovascular disease^d^, No. (%) |  |  | 0.564 |
| yes | 112 (5.1) | 25 (4.6) |  |

Abbreviations: BMI, body mass index; BDI, Beck Depression Index; LDL, low density lipoprotein cholesterol; HDL, high density lipoprotein cholesterol; HTN, hypertension; DM, diabetes; CVD, cardiovascular disease.

a Study participants who included in the final analyses.

b Study participants who excluded for the final analyses.

c Continuous variables are age, BMI, BDI, LDL, and HDL. Age, BMI, and LDL were described as mean (SD). HDL and BDI were described as median (IQR).

d Categorical variables are sex, education, drinking and smoking status, exercise, HTN, DM, and CVD status. All categorical variables were described as No. (%).
